# Smoking Impairs Women’s Verbal Learning and Memory Performance More than Men’s: An International Web-Cohort Study of 70,000 Participants

**DOI:** 10.1101/2020.08.25.20181735

**Authors:** C.R. Lewis, J.S. Talboom, M.D. De Both, A.M. Schmidt, M.A. Naymik, A.K. Håberg, T. Rundek, B.E. Levin, S. Hoscheidt, Y. Bolla, R.D. Brinton, M. Hay, C.A. Barnes, E. Glisky, L. Ryan, M.J. Huentelman

## Abstract

**Background:** Vascular contributions to cognitive impairment and dementia (VCID) include structural and functional blood vessel injuries linked to poor neurocognitive outcomes. Smoking might indirectly increase the likelihood of cognitive impairment by exacerbating the risks associated with underlying vascular disease. Sex disparities in VCID have been reported, however, few studies have assessed the sex-specific impact of smoking on cognitive function and with contradictory results. This is an important topic since smoking and cardiovascular disease negatively impact health and possibly women have the greater lifetime risk of stroke and dementia than men. In this study, we sought to investigate the effect-modification of sex on the relationship between smoking, cardiovascular disease and verbal learning and memory function.

**Methods:** Using MindCrowd, a web-based cohort of over 70,000 people aged 18 - 85, we investigated whether sex modifies the impact of smoking and cardiovascular disease on verbal memory performance on a paired-associate learning task using both multiple regression and propensity matching approaches. Artificial error introduction and permutation testing underscored the stability of our results. To demonstrate the necessity of large sample sizes to detect an interaction of sex and smoking, we performed down sampling analyses.

**Findings:** We found significant interactions in that smoking impacts verbal learning performance more in women and cardiovascular disease more in men across a wide age range.

**Interpretation:** These results suggest that smoking and cardiovascular disease impact verbal learning and memory throughout adulthood. Smoking particularly affects learning and memory in women and cardiovascular disease has a larger effect in men. Although the reasons for these sex-modification effects are not entirely understood, our findings highlight the importance of considering biological sex in VCID.

**Funding:** Mueller Family Charitable Trust; Arizona Alzheimer’s Consortium; Flinn Foundation; The McKnight Brain Research Foundation; NIH-NIA grant R01-AG049465.

## INTRODUCTION

Vascular contributions to cognitive impairment and dementia (VCID) include blood vessel injuries that can cause significant changes to memory, thinking, and behavior (1). Vascular diseases are also associated with increased risk for Alzheimer’s disease (AD) (2), which is the 6th leading cause of death in the US with increasing numbers and financial tolls (3,4). Vascular-related dementia is the second most common cause of cognitive decline, with only AD-related dementia being more prevalent (5). Several extensive cohort-based studies have found that cardiovascular disease risk factors such as tobacco smoking are associated with cognitive decline and increased risk of dementia in older age (6).

Despite early reports suggesting that smoking tobacco might have beneficial effects on cognition (7,8) and a reduced risk of dementia (9), more recent evidence clearly suggests that active smoking has neurotoxic effects on the brain (10–12) and is associated with a doubling of dementia risk for older adults (13). Smoking tobacco might indirectly increase risk of cognitive impairment by exacerbating the subclinical risks associated with underlying vascular disease. Smoking causes vascular damage, including carotid artery disease, atherosclerotic plaque formation, increased platelet aggregation, compromised endothelial cell function, arterial stiffness and increased systolic blood pressure, which all contribute to stroke risk (14–16). The presence of smoking with hypertension is one of the greatest risk factors for acute myocardial infarction and stroke, according to a meta-analysis of 10 cohort studies which included more than 60,000 people (17). Tobacco smoking is also a major risk factor for cardiovascular disease and chronic obstructive pulmonary disease (COPD), both of which can decrease cognitive function (18). Importantly, prevalence rates of any dementia and AD may be greater in women than men (19–21). A recent study found that women had a stronger association between vascular risk factors and worse cognition than men in a middle-aged Hispanic/Latino population (22). However, it is currently unknown if the effects of smoking on verbal learning and memory function are different between men and women.

Several studies have found an association between smoking, cognitive decline, and dementia (23–25). Nevertheless, the majority of these studies do not assess biological sex as a main variable of interest but instead use sex as a covariate. As a covariate, the variance due to sex is only being controlled for and not directly explored. Thus, the question whether smoking affects cognition in men and women differently remains unresolved. One methodological issue that might explain why so few studies have directly examined this important question pertains to the need for relatively large sample sizes to be adequately powered for interaction, or moderation, analyses. It is estimated that detecting an interaction requires at least four times the sample size than needed to detect a main effect (26,27).

The few studies that assess a smoking by sex interaction or test women and men separately, have typically been underpowered and have found contradicting results. Some studies suggest no sex effect or that smoking may affect men’s cognition more strongly than women’s (13,25,28–30); however, other studies have found a larger effect of smoking on cognition in women (31,32). It does appear, however, that smoking often impacts women’s risk to a greater extent for diseases that occur outside of the central nervous system. A meta-analysis of more than 2.4 million people suggests that, compared with nonsmokers, women who smoke have a 25% greater relative risk of coronary heart disease than men who smoke, independent of other cardiovascular risk factors (33). Women smokers may also have a greater relative risk of lung cancer than men who smoke (34). Others have found the effects of smoking in various disease models such as bone fragility and Crohn’s disease differ between men and women (35,36). Furthermore, there appears to be a sex difference in the behavioral response to the acute effect of smoking in prepulse inhibition, a measure of startle reflex (37). Importantly, human and animal studies point to many structural and functional sex differences in nicotinic acetylcholinergic brain physiology (for review see (38)). Collectively, these data suggest sex-specific effects should be considered a high priority of inquiry when assessing the relationship between smoking status and cognition.

The primary objective of this study was to determine whether biological sex moderates the impact of smoking on memory performance in healthy adults aged 18 – 85 years old. We performed down sampling analyses to estimate the minimum sample size required to detect this interaction. In addition, given the known deleterious effects of smoking on the cardiovascular system and the established relationship between cardiovascular disease and cognitive impairment and dementia (39–42), we further tested if sex moderates the combined influence of diabetes, heart disease, hypertension, and stroke history on memory performance. We hypothesize that both smoking and an integrated cardiovascular disease index would have greater detrimental effects on paired-associate learning in women compared to men. For this study, we leveraged MindCrowd (www.mindcrowd.org), a web-based cohort of more than 70,000 persons aged 18 to 85 years from which memory and demographic data were collected.

## METHODS

### Study participants

In January of 2013 we launched our study site at www.mindcrowd.org. Website visitors, who were 18 years or older, were asked to consent to our study before any data collection via an electronic consent form. Approval for this study was obtained from the Western Institutional Review Board (WIRB study number 1129241).

As of March 17, 2020 MindCrowd has recruited 84,260 qualified participants from around the world aged 18 – 85 with 64.3% women and 35.7% men. An overrepresentation of women has been previously described in studies drawn from the general population (43). Across the entire sample, 7.9% of participants reported being a current smoker (Figure 1). The entire demographic, health, lifestyle, and medical variable composition of our study cohort can be found in Supplemental Table 1.

**Figure 1:**
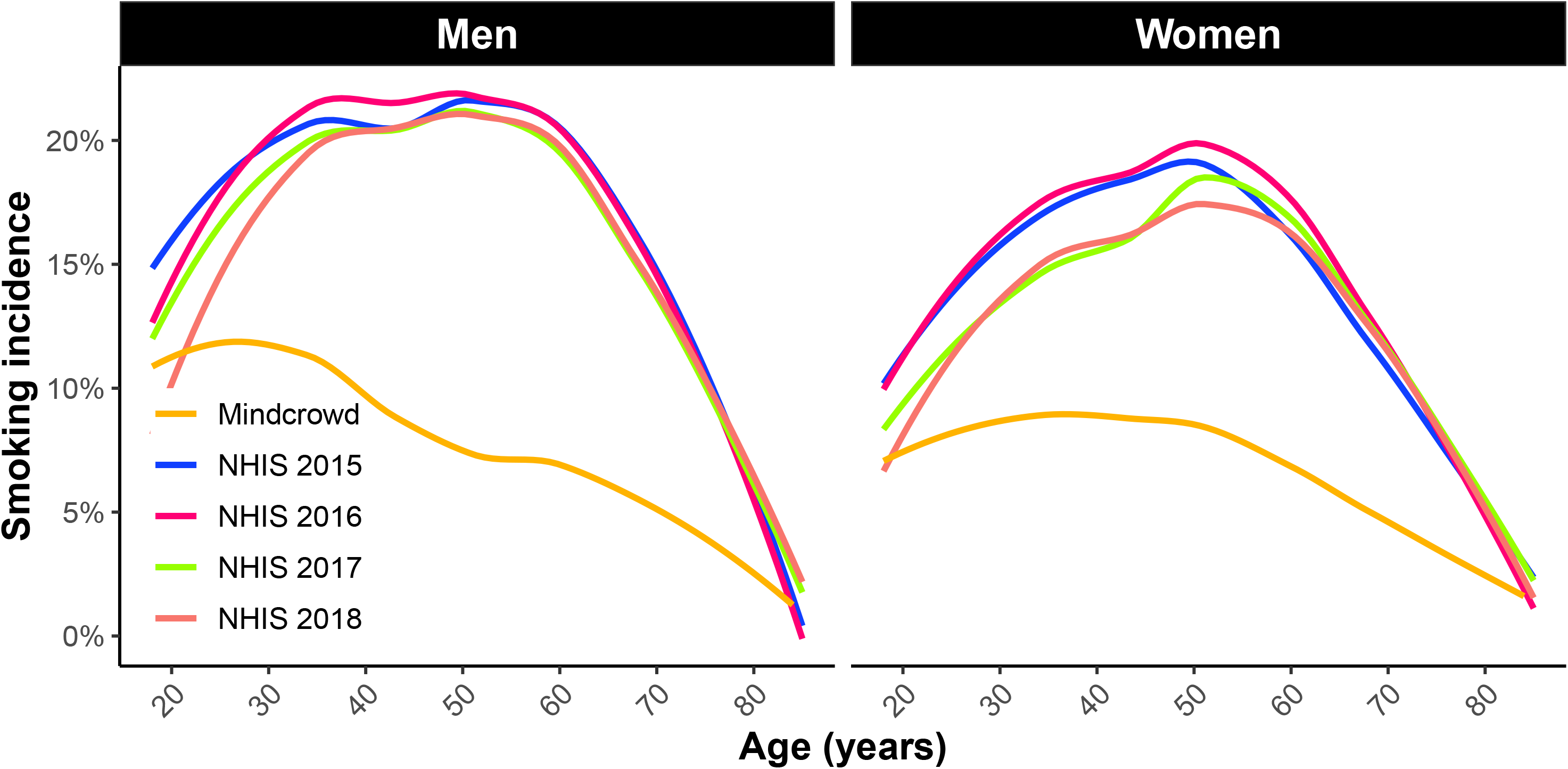
A visual representation of US smoking rates by age derived from data from the National Health Interview Study (NHIS) for years 2015-2018 and MindCrowd smoking rates per age for men (A) and women (B). The NHIS is is one of the major household survey-based data collection programs of the National Center for Health Statistics (NCHS) which is part of the Centers for Disease Control and Prevention (CDC). These data demonstrate that men typically have higher smoking rates compared to women of the same age and that MindCrowd, while demonstrating lower rates of smoking overall, follows the same trend as observed by the NHIS.

After consenting to the study and answering five demographic questions (age, sex, years of education, primary language, and country of residence), participants completed a web-based paired-associates learning (PAL) task. For this cognitive task, during the learning phase, participants were presented 12 word-pairs, one word-pair at a time (2 sec/word-pair). During the recall phase, participants were presented with the first word of each pair and were asked to use their keyboard to type (i.e., recall) the missing word. This learning-recall procedure was repeated for two additional trials. Before beginning the task, each participant received one practice trial consisting of three word-pairs not contained in the 12 used during the test. Word-pairs were presented in different random orders during each learning and each recall phase. The same word pairs and orders of presentation were used for all participants. The dependent variable/criterion was the total number of correct word pairs entered across the three trials (i.e., 36 is a perfect score). Upon completing the PAL task, participants were directed to a webpage asking them to fill out an additional 17 demographic and health/disease risk factor questions including if they are a current smoker. Other questions included: marital status, handedness, race, ethnicity, number of daily prescription medications, a first-degree family history of dementia, and yes/no responses to the following: seizures, dizzy spells, loss of consciousness (more than 10 min), high blood pressure, diabetes, heart disease, cancer, stroke, alcohol/drug abuse, brain disease, and memory problems. Next, participants were shown their results and were provided with different comparisons to other test takers based on the average scores across all participants, as well as across sex, age, education, etc. On the same page, participants were also provided with the option to provide contact information if they wanted to be recontacted for future research.

### Statistical Analyses

#### Multivariate Linear Regression

To investigate the sex x smoking interaction on memory performance we ran a linear model (LM) controlling for various health and lifestyle factors from self-report including: age, race, ethnicity, marital status, handedness, education attainment, number of daily medications, history of diabetes, seizures, cancer, stroke, hypertension, heart disease, family history of Alzheimer disease, drug abuse, loss of consciousness, and dizziness. The dependent variable was the total number of correct word pairs entered across the three trials of PAL tests (range of 0 - 36). We report standardized beta coefficients, standard error (SE), and *p* values.

To assess the interaction between cardiovascular disease x sex on PAL performance, we created a cardiovascular disease composite score by computing a sum number of CVD factors including heart disease, hypertension, diabetes, and stroke history (range of 0 – 4). Individuals were treated as groups based on their composite score (0 as the control group compared to 1, 2, and 3+ of these conditions). Participants with a sore of 3 or 4 were combined into one group to create an adequate sample size. We ran a linear model (LM) controlling for age and education level. We report standardized beta coefficients, standard error (SE), and *p* values.

#### Propensity Score Matching

In addition to running a multivariate linear regression model, we also conducted propensity score matching. Propensity score matching (PSM) has the benefit of reducing bias and variance (44); however, this is often achieved at the expense of sample size compared to regression analysis. We matched participants on all variables listed as covariates in the multivariate linear model. Matching was performed using the R package, MatchIt (version 3.0.2). Effect sizes were estimated from the matched cohort using the R package, Zelig (version 5.1.6.1). Due to constraints on sample sizes, we conducted the PSM model collapsed across all ages on men and women separately. Zelig uses least squares regression on matched data to estimate the partial effect on an outcome of interest, in our case, total word pairs correct (45,46). We also report effect sizes with credible intervals. The credible interval is the Bayesian version of a confidence interval and can be interpreted as a probability (i.e., there is a 95% probability that the effect size is between X and Y word pairs).

#### Down Sampling

In order to demonstrate the importance of our large sample size for generating reliable effects, we ran 1000 down-sample linear regression models of the MindCrowd cohort between the ages of 18 and 85 years for the interaction effect of smoking x sex on paired-associate learning (PAL) for each indicated total sample size. Total sample sizes ranged from 268 to 13,400 and each group (smoker and non-smoker and sex) was sampled at equal size per age 18-85.

#### Artificial Error Introduction

To investigate the potential role false-report error may play on the smoking effect, we used a Monte Carlo simulation to determine the effect of introducing artificial error (in addition to any real self-report error already in the data). The additional error was added by randomly re-assigning the self-reported smoking status (smoker or non-smoker) to various percentages of the cohort (stepwise from 1–10% of individuals) and re-analyzing the effect of smoking using our complete statistical model. This process was performed a total of 10,000 times for each error percentage, and the resulting influences on the p-value are reported using boxplots.

#### Permutations

We examined the smoking effect on PAL through the use of permutation testing. Permutation testing is an approach utilized to determine the probability of a false-positive finding if the null hypothesis were true. To create the permuted datasets, we randomly assigned the smoking status for each participant before analyzing the complete model in the whole cohort, women only, and men only. We also ran permutation tests on the interaction between sex x smoking on PAL. This process was performed one million times per model.

## RESULTS

### Multivariate Linear Regression

#### Smoking

In women and men 18–85 years old our linear model (LM) revealed a significant omnibus effect of smoking [*F*(30, 81670) = 560. 58, *p* = 0e+00; Figure 2]. The model also revealed a significant sex x smoking interaction [β = 0.99, SE = .22, *p* = 4e-6]. Simple effects analyses revealed that women’s memory is negatively impacted by smoking [β = -1.01 word pairs, SE = .14, *p* = 7.83e-13; Figure 3] whereas men’s memory was not significantly impacted [β = -0.27 word pairs, SE = .18, *p* = 0.135: Figure 3].

**Figure 2.**
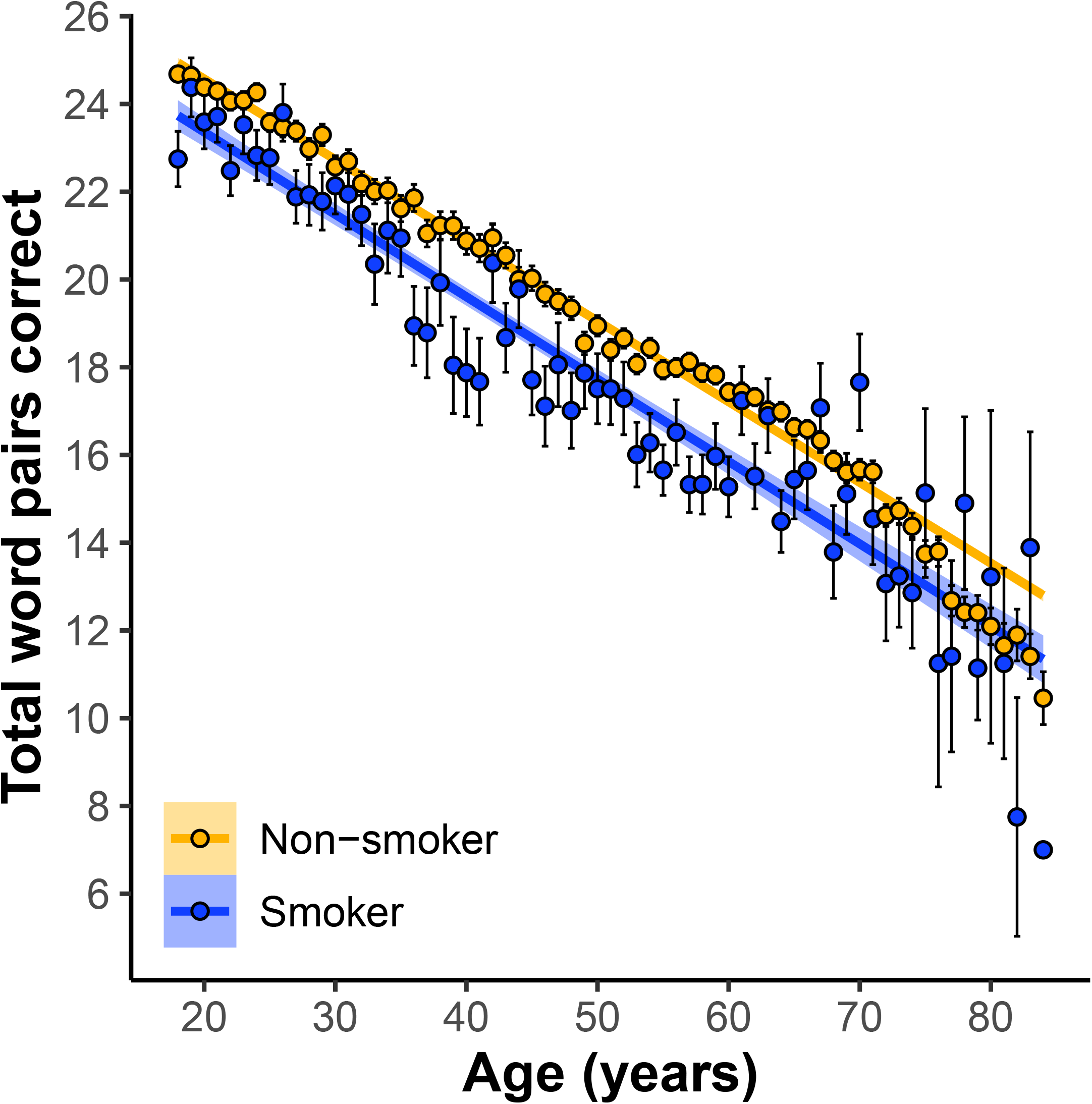
Participants who self-report current smoker status perform worse on paired-associates learning (PAL) compared to non-smokers across 18-85 year olds. Linear regression fit (line fill ±95% confidence interval [CI]) of the PAL total number of correct from 18 to 85 years old. Lines were split by Smoker versus Non-smoker. [F(30, 81670) = 560. 58, p = 0e+00, N = 81, 700]

**Figure 3.**
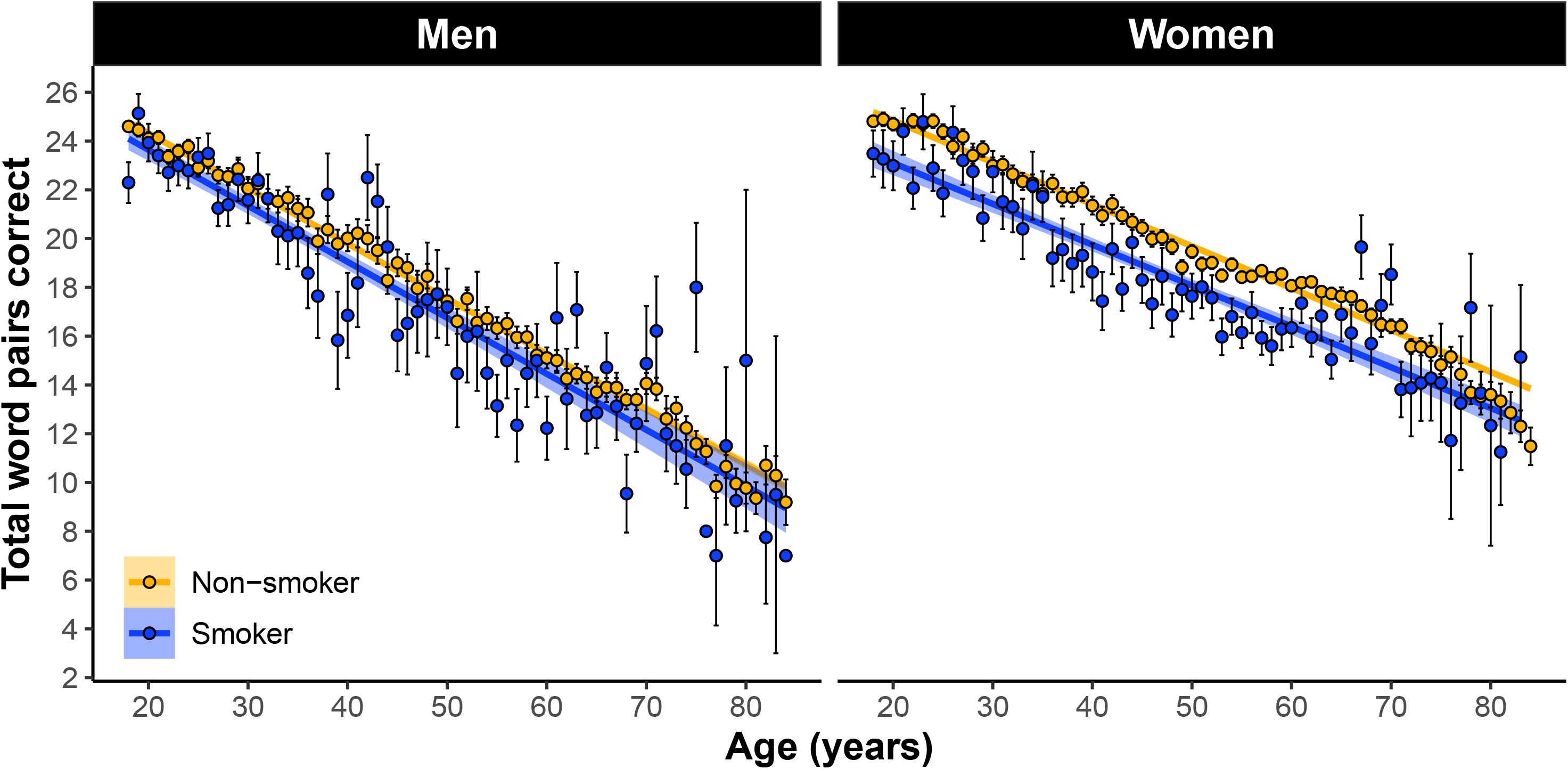
The main effect of self-reported smoking status on PAL performance across 18-85 year olds separately between (A) men and (B) women. Simple effects analyses revealed that women’s memory is negatively impacted by smoking [β = -1.01 word pairs, std error = .14, p = 7.83e-13; Panel B] whereas men’s is not significantly impacted [β = -0.27 word pairs, std error = .18, p = 0.135: Panel A]. Shown are the linear regression fit lines (±95% confidence interval [CI]) of the PAL total number of correct from 18 to 85 years old.

#### Cardiovascular disease

In the whole cohort, 77.4% of women and 78.1% of men had a CVD sum of 0, 17.9% of women and 16.6% of men had a CVD sum of 1, 4.1% of women and 4.3% of men had a CVD sum of 2, and 0.6% of women and 1% of men had a CVD sum score of 3+. In women and men 18–85 years old our linear model revealed a significant omnibus effect of cardiovascular disease [*F*(31, 81669) = 542.88, *p* = 0e+00: Figure 4]. The model also revealed a significant sex x cardiovascular disease interaction in the 1 group [β = -0.84 word pairs, SE = 0.14, *p* = 6.41e-9], 2 group [β = -1.38 word pairs, SE = 0.27, *p* = 2.08e-7], and 3+ group [β = -1.64 word pairs, SE = 0.54, *p* = 0.002]. Simple effects analyses revealed the negative impact of cardiovascular disease on memory had slightly larger effect sizes in men compared to women in the 1 group [men: β = -0.64 word pairs, SE = 0.12, *p* = 1.22e-7; women: β = -0.41 word pairs, SE = 0.09, *p* = 3.74e-6], the 2 group [men: β = -0.99 word pairs, SE = 0.23, *p* = 1.2e-5; women: β = -0.67 word pairs, SE = 0.16, *p* = 3.18e-5], and the 3 group [men: β = -1.80 word pairs, SE = 0.41, *p* = 1.3e-5; women: β = -1.27 word pairs, SE = 0.35, *p* = 0.0003; Figure 5].

**Figure 4.**
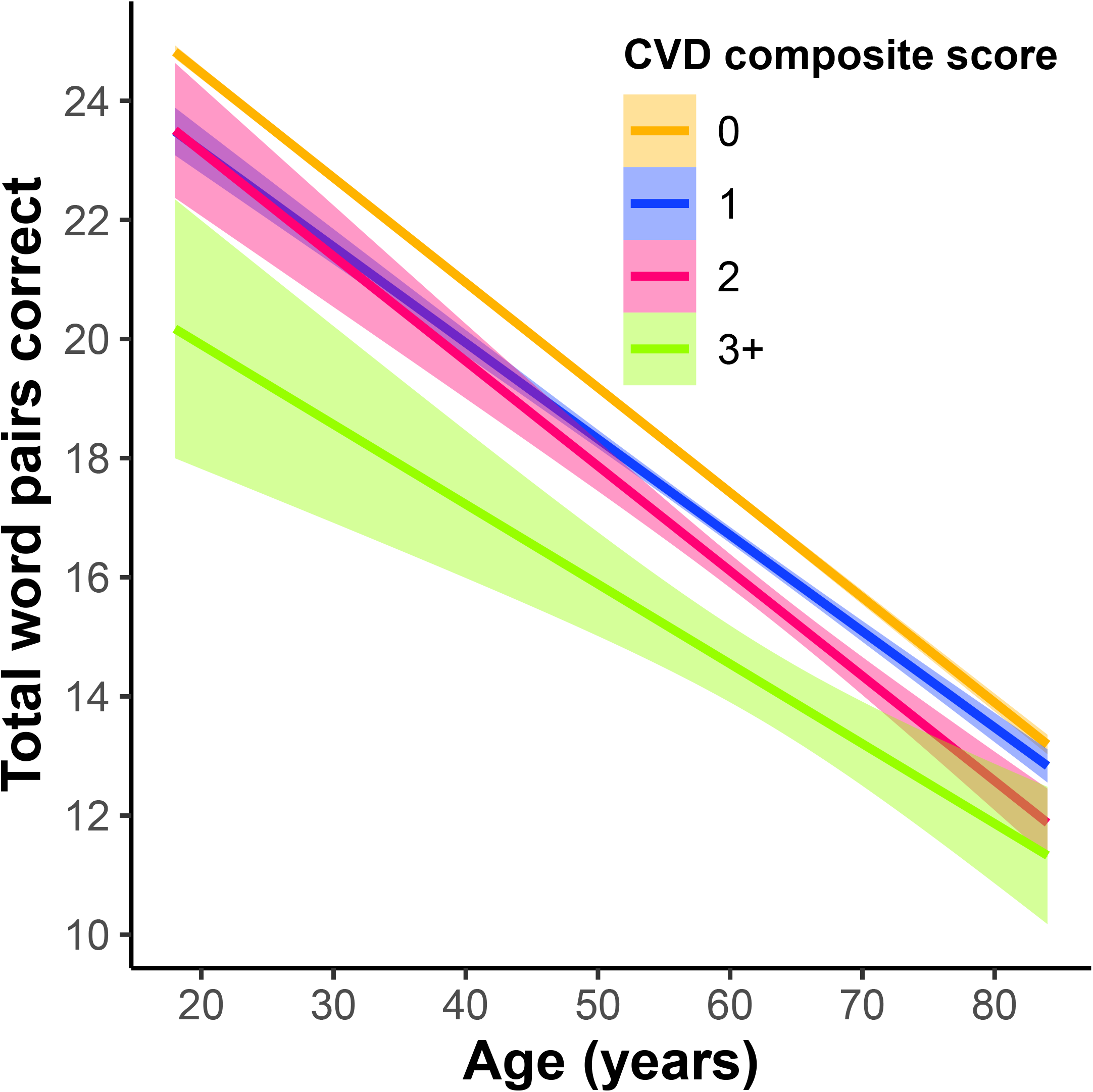
The main effect of a self-reported cardiovascular disease (CVD) composite score on PAL performance across 18-85 year olds. CVD composite was a sum score of self-reported heart disease, hypertension, diabetes, and stroke. Individuals were treated as groups based on their composite score (0 as the control group compared to 1, 2, and 3+). Linear regression fit (line fill ±95% confidence interval [CI]) of the PAL total number of correct from 18 to 85 years old [F(31, 81669) = 542.88, p = 0e+00].

**Figure 5.**
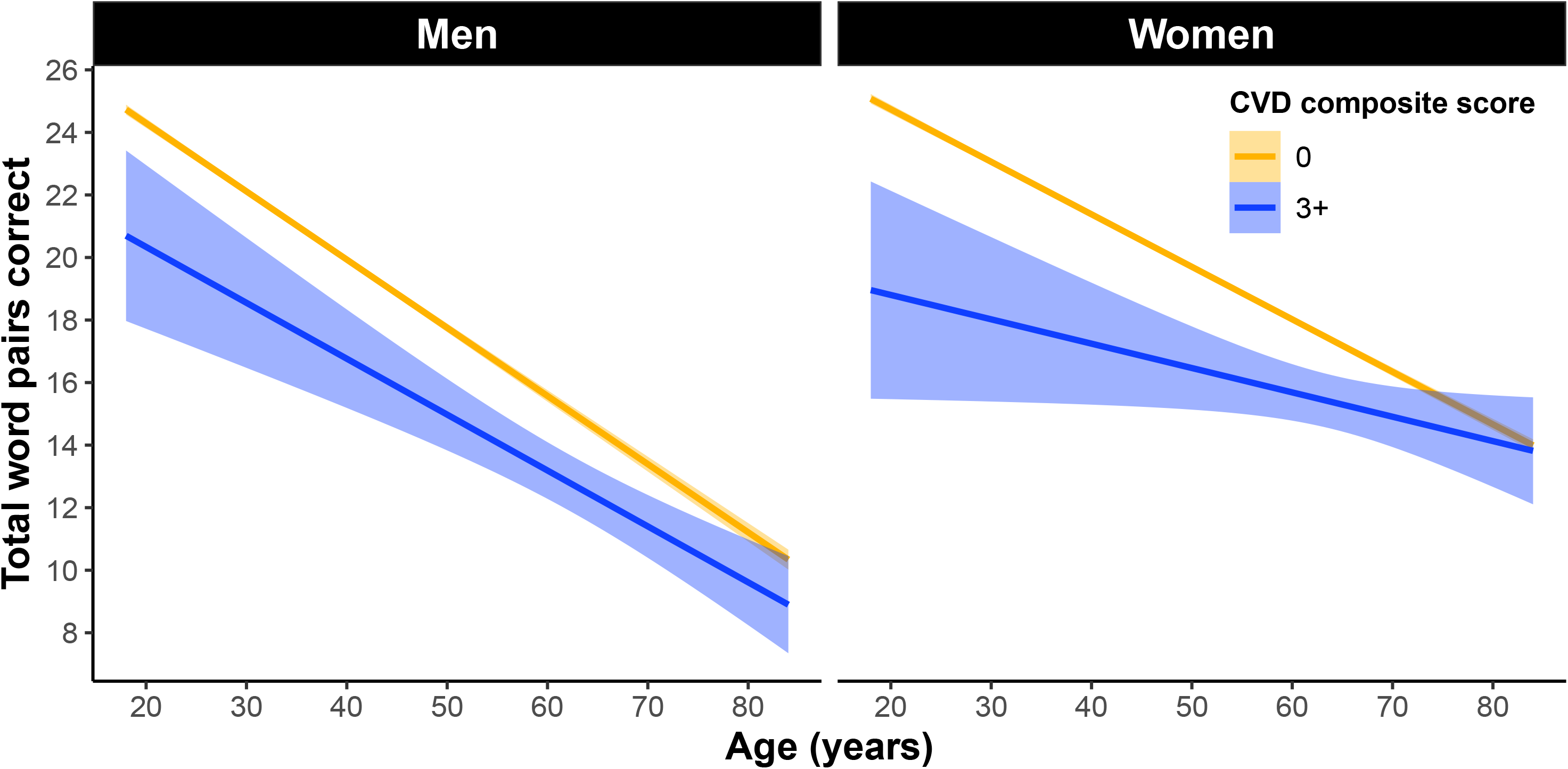
The main effect of a self-reported CVD composite score on PAL performance across 18-85 year olds separately between men and women. Simple effects analyses revealed the negative impact of cardiovascular disease on memory had slightly larger effect sizes in men compared to women in the 3 group [Panel A men: β = -1.80 word pairs, std error = 0.41, *p* = 1.3e-5; Panel B women: β = -1.27 word pairs, std error = 0.35, *p* = 0.0003].

### Propensity Score Matching

When collapsed across age, PSM suggested there is no effect of smoking on memory in men [β = -0.21 (−0.62 to 0.19, 95% credible interval)] and a negative effect in women [β = -0.54 (−0.14 to -0.93, 95% credible interval)] (Figure 6).

**Figure 6.**
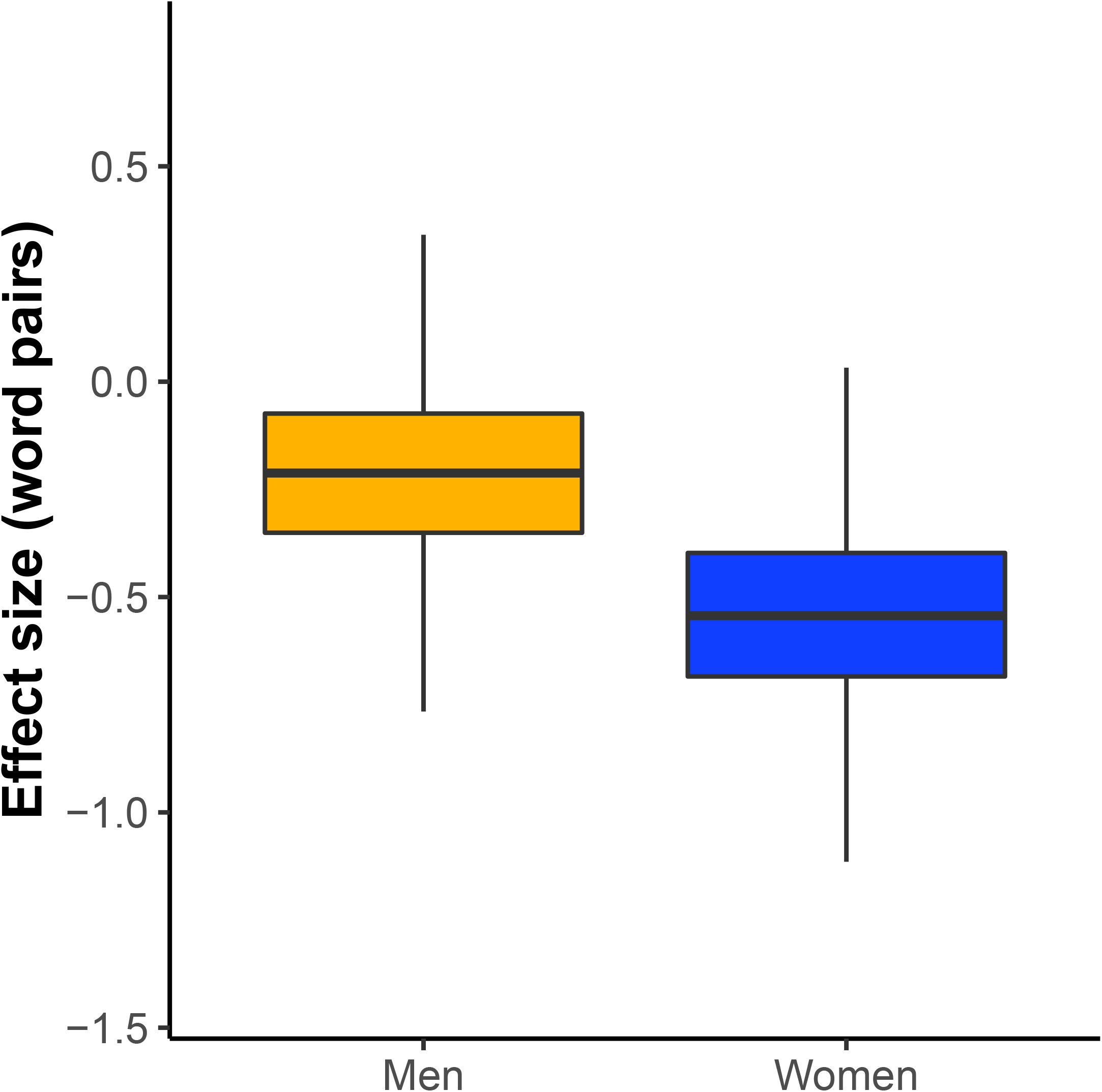
The propensity score matching main effect of self-reported smoking on PAL performance across all ages separately between men and women. We conducted propensity score matching (PSM) analysis matching smokers and non-smokers for various health and lifestyle factors from self-report including: age, race, ethnicity, marital status, handedness, education level, number of daily medications, history of diabetes, seizures, cancer, stroke, hypertension, heart disease, family history of Alzheimer disease, drug abuse, loss of consciousness, and dizziness. The dependent variable was the total number of correct word pairs entered across the three trials of PAL tests (range of 0 - 36). When collapsed across age, PSM suggested there is no effect of smoking on memory in males [β = -0.21 (−0.62 to 0.19, 95% credible interval)] and a negative effect in females [β = -0.54 (−0.14 to -0.93, 95% credible interval)].

### Down Sampling

The horizontal red line in Figure 7A indicates the effect size estimated by the largest-sized sample. Green filled circles indicate an individual down-sampled comparison that resulted in a statistically significant association (*p* <0.05). We performed 1,000 of these simulations at each down-sampled size. At a cohort size of approximately 10,000 samples, 50% of down-sampled comparisons resulted in the observation of a significant smoking x sex interaction (Figure 7B). Even at the largest sample size shown, there are still some observed down-sampled comparisons that result in a non-significant association (black circles). These data illustrate why there is a concern about small sample sizes and their ability to result in misestimated β values and the probability of type 2 errors. Additionally, note that at sample sizes below 6000 it is possible to observe significant associations in the opposite direction of the actual effect (e.g. smoking enhances performance).

**Figure 7:**
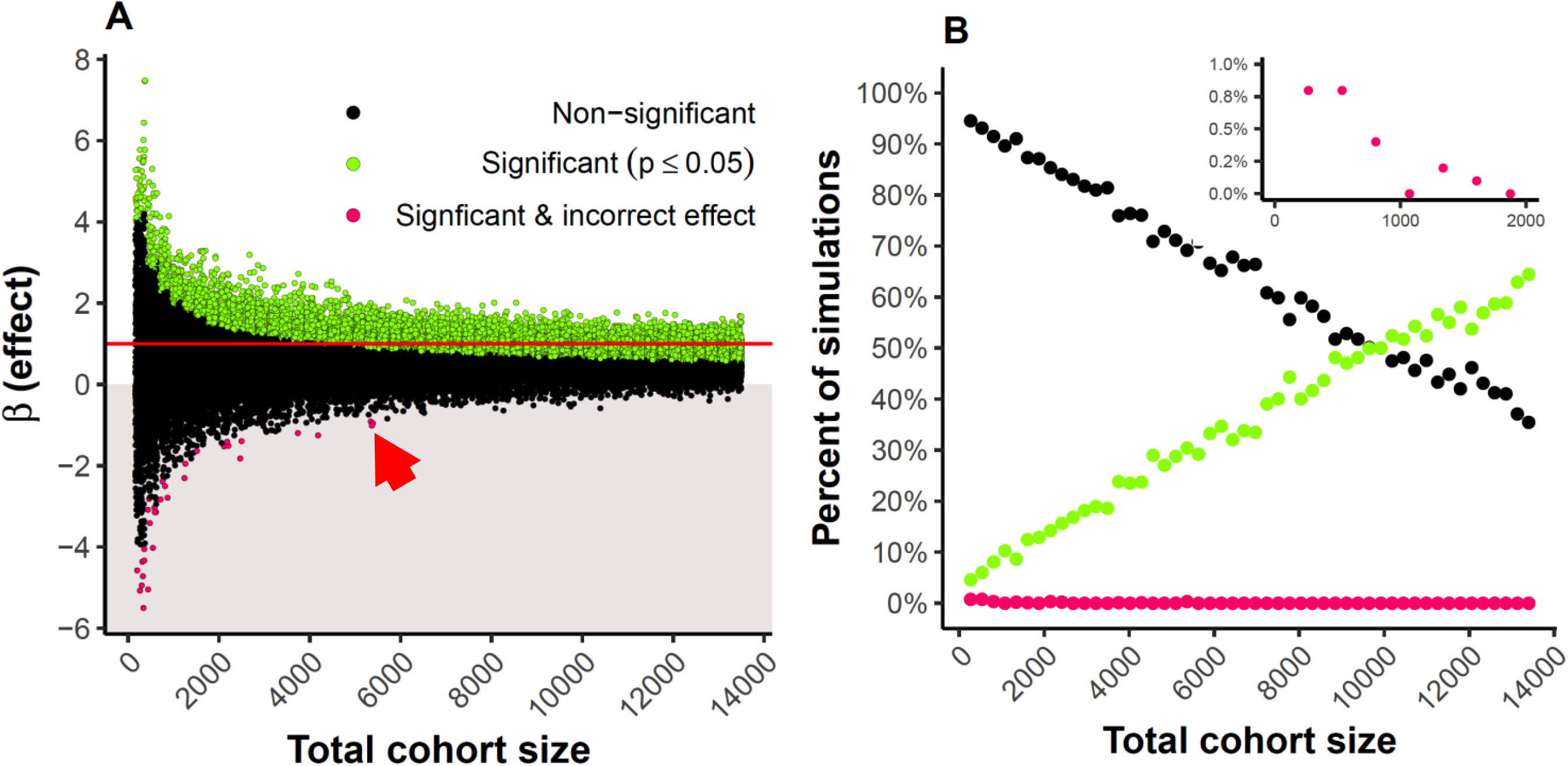
**(A)** We conducted 1000 down-sample linear regression models of the MindCrowd cohort between the ages of 18 and 85 years for the interaction effect (β, y axis) of sex x smoking on paired associate learning (PAL) for each indicated total sample size (x axis). For each analysis, we had an equal amount of smokers and non-smokers and women and men. The horizontal red line indicates the effect size estimated by the total study sample. Green filled circles indicate an individual down-sampled comparison that resulted in a statistically significant association (*p* <0.05), black dots are non-significant comparisons. Red arrow highlights that at samples sizes approximating 5000 one could potentially produce a significant beta value with the opposite sign from the largest sampled model. **(B)** Panel B is the same data displayed to easily see the positive relationship between significant betas and sample size.

### Artificial Error Introduction

A Monte Carlo simulation (Figure 8) was used to determine the effect of introducing artificial error (in addition to any real self-report error already in the data). The error was introduced in 1% increments (x-axis) by randomizing smoker status. Each box represents 10,000 model simulations, and plotted are the p-values for each simulation. The red line represents a significance level of α = 0.05. At 0% simulated error, we report our measured *p*-value. In the whole cohort (Figure 8A) and women specifically (Figure 8B), we measured a significant effect in > 90% of simulations after 10% additional self-report error was introduced. For the sex x smoking interaction term, 75% of simulations were statistically significant after 10% additional self-report error was introduced (Figure 8C).

**Figure 8.**
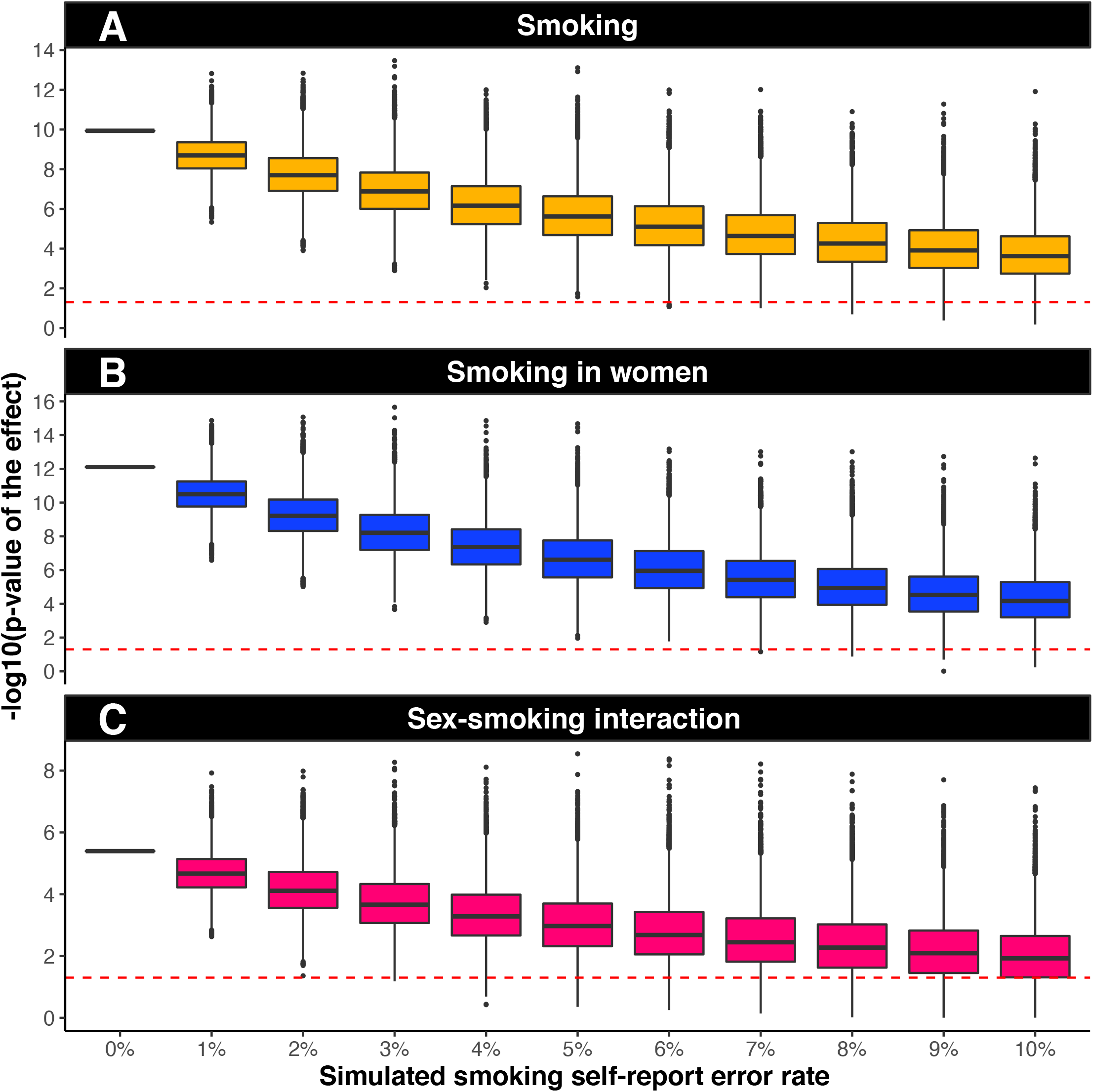
Artificial error introduction suggests the present smoking results are likely not due to self-report error. A Monte Carlo simulation was used to determine the effect of introducing artificial error (in addition to any real self-report error already in the data). Error was introduced in 1% increments (x-axis) by randomizing smoker status. Each box represents 10,000 model simulations, and plotted are the p-values for each simulation. The red dashed line represents a significance level of α = 0.05. At 0% simulated error, we report our measured p-value. In the whole cohort and women specifically (panels A & B), we measure a significant effect in > 90% of simulations after 10% additional self-report error is introduced. For the sex x smoking interaction term, 75% of simulations are statistically significant after 10% additional self-report error is introduced.

### Permutations

From one million permutations performed for the main effect of smoking in the whole cohort (Figure 9.A) and the main effect of smoking in women only (Figure 9.B), not a single *t*-statistic was observed to be more extreme than our reported *t*-statistic. This suggests that the odds of our findings being observed due to chance alone is less than one in a million. However, our reported *t*-value for the men only analysis did overlap with values calculated in the permutation tests demonstrating confidence in our non-significant finding (Figure 9.C). Further, we ran the same permutation tests on the sex x cardiovascular interaction term and showed that our full model *t*-value did not overlap with any permutation test (Figure 9.D).

**Figure 9.**
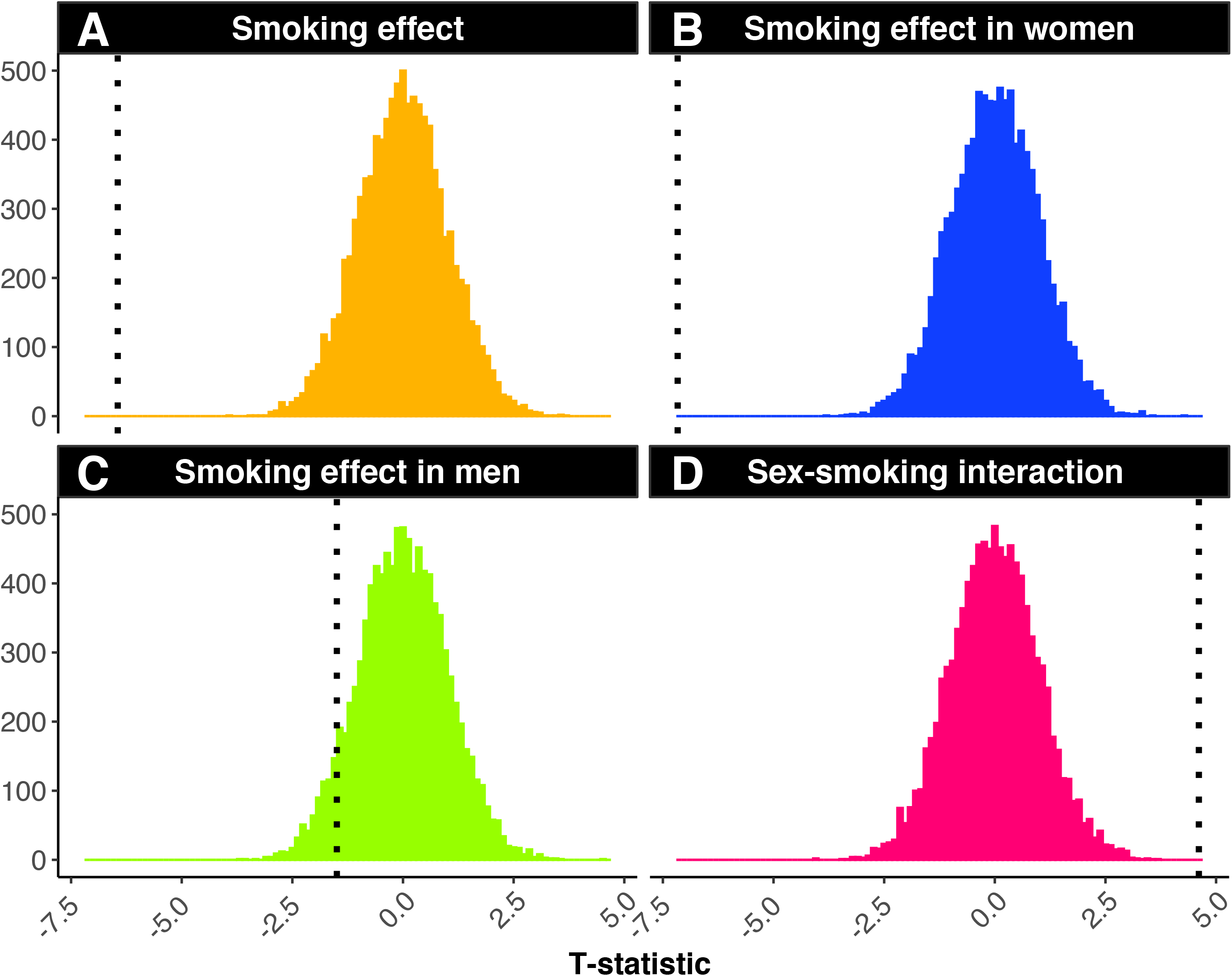
We examined the smoking effect on PAL through the use of permutation testing. This was performed one million times per model. The smoking data label for every participant was randomly assigned and the t-statistic for the main effect of smoking in the whole cohort (A), women only (B), and men only was re-calculated (C). We also conducted permutation tests on the interaction between sex x smoking on PAL (D). Black dashed line indicates the full model results statistic. Results from permutation testing indicate the present results are likely not due to chance.

## DISCUSSION

In this large web–based study, we found current cigarette smoking and cardiovascular disease are associated with worse memory performance in adults as young as 18 years. Furthermore, we found significant sex-modification of these associations showing that the impact of smoking on verbal recall was worse in women whereas the impact of cardiovascular disease on memory performance was worse in men. These findings are important because according to the U.S. Department of Health and Human Services, cigarette smoking is the leading cause of preventable disease and death in the United States and accounts for about 1 in every 5 deaths (47). In 2018, nearly 14 of every 100 U.S. adults aged 18 years or older (13.7%) smoked cigarettes, which translates to about 34.2 million adult smokers (48). In addition, cardiovascular disease is the leading cause of morbidity and mortality worldwide, and an important predictor of cognitive decline and VCID.

We found that sex modifies the relationship between smoking and verbal recall in that women are negatively affected to a larger degree than men. This finding is in agreement with several studies that reported a larger effect of smoking on cognition in women (31,32), but stands in contrast with other smaller studies that found no sex effect or that smoking might affect cognition more strongly in men (13,25,28–30). These results also align well with studies that suggest smoking impacts coronary heart disease and lung cancer more in women than in men (33,34). We used down sampling analyses to demonstrate the importance of large sample sizes to ensure reproducible interaction effects of our model. These analyses suggest that a study sample of at least 10,000 is needed to observe a significant sex by smoking interaction at least 50% of the time. This finding highlights the possibility that many previous studies may have been underpowered to find the interaction.

In the United States, men (15.6%) were more likely to be current cigarette smokers than women (12.0%) according to a 2018 report (48) and we replicated this finding within our MindCrowd cohort (Figure 1). In addition, women on average smoke fewer cigarettes per day and have lower salivary cotinine levels compared with men. However, smoking rates for women have increased relative to smoking rates for men in the US and the popularity of smoking in women from low to middle-income countries may be increasing (49,50). This is of particular interest since there are higher numbers of Alzheimer’s disease cases in women than men (19–21) and these data suggest that smoking could potentially accelerate these trends. While men may be at a slightly higher risk for VCID throughout most of the lifespan, some risk factors for VCID more adversely affect women such as preeclampsia, menopause, and hormone replacement (51). Not only does the risk of dementia increase with age, but normative decreases in many cognitive abilities occur across the lifespan (52), therefore a better understanding of modifiable contributors, such as smoking, to cognitive function is essential.

In addition to sex differences in smoking behaviors, sex differences in the cholinergic system are possible biological mechanistic explanations for why smoking may have a more substantial impact on cognition in women compared to men. Animal studies have shown both nicotine-exposed and non-nicotine exposed female rats exhibit higher cholinergic receptor (NAChR) densities than their male counterparts (53). Since NAChR innervation influences several cognitive functions and neurotransmitters (54), perhaps higher expression in women exacerbates the effects of smoking on cognition. However, it is not clear whether smoking-related changes in cognition are primarily due to nicotine exposure or the complex chemical makeup of cigarette tobacco and its additives (47). This is becoming an important distinction as the prevalence of adult e-cigarette use/vaping increased from 2.8% in 2017 to 3.2% in 2018 (48). However, since we did not ask about vaping specifically, further research is needed to monitor the relationship between e-cigarette use and memory performance since it is possible that the effects of e-cigarette use will differ compared to smoking tobacco due to the differing mix of chemical exposures.

Given the established relationships between smoking and cardiovascular disease, we also tested whether sex moderates the impact of cardiovascular disease on memory performance. Although much of the prior research in this domain has focused on cardiovascular risk scores (for review see (6)), we used number of CVD disease incidents as opposed to risk factors. Nevertheless, we can draw some comparisons between these investigations. The existing cross-sectional associations between cardiovascular disease risk and cognitive function in the literature are largely consistent with results obtained in this study between cardiovascular disease and memory performance. However, comparison with these findings for the sex effect is limited because of differences in study analyses. As with smoking studies, the majority of this research investigates sex as a covariate in the primary analyses instead of comparing men and women separately or investigating an interaction (for review see (6)). We found that sex moderates the relationship between cardiovascular disease and cognitive performance in that men are affected to a larger degree. However, the few studies that investigated men and women separately have found women’s cognitive performance to be slightly more impacted. For example, Kaffashian et al. (2011) investigated the Framingham cardiovascular risk profile and cognitive function and 10-year decline separately between men and women and reported larger effects in women across their cognitive batteries in 35-55 year-olds (55). In a cohort of community-dwelling adults without clinical heart disease, the Framingham Cardiac Risk Score (FCRS) was associated with the rate of cognitive decline in women, but not men (56). Yet another cohort of Mexican Americans demonstrated that higher predicted cardiovascular disease risk was associated with greater change in errors on multiple cognitive tests and that these associations were larger and more significant in women than men (57). Lastly, a recent study found women had a higher association between vascular risk factors and worse cognition compared to men in a middle-aged Hispanic/Latino population (22). It is unclear why we found larger effects in men, however, it could be due to the broad age range included in our study. The relationship between cardiovascular disease and cognitive function is primarily studied in older adults, with a few examples including 35 and older (42,55,58,59), and one study including 18-30 year-olds (60). Understanding the relationship between cardiovascular health and cognitive function in young adulthood may be necessary for understanding possible treatment and intervention opportunities.

Due to the large, widely available, and electronic nature of our study cohort, we rely on self-report answers to demographic, lifestyle, and health questions (61). Current studies comparing self-report data given over the Internet versus data collected in-person show anywhere from a 0.3–20% discrepancy for height and weight measurements (62,63). For some socially unacceptable measures (like smoking) internet self-report may actually have higher accuracy since the pressure to “perform” well in the presence of an investigator is removed when answering questions electronically (64). To investigate the potential role that false-report error may play on our smoking effect, we re-analyzed the smoking effect after introducing additional error into the smoking self-report response. The additional error was added by randomly flipping the smoking response to various percentages of the cohort (stepwise from 1–10% of individuals) and re-analyzing the effect of smoking using our complete statistical model a total of 10,000 times for each error percentage. These results suggest that it is unlikely that smoking self-report error is driving our results. Lastly, PAL was tested cross-sectionally in the cohort; therefore, determinations about the influence of collected factors on trajectories of change in performance across time within an individual subject are not possible. Additional longitudinal-based studies will be necessary to identify this class of variables.

There are limitations of this study to acknowledge. First, the smoking rate in MindCrowd is lower than national averages which may reflect the tendency of healthier people to participate in observational research (65). However, since we used propensity matching to confirm the sex effect, this suggests our results are generalizable to the broader population. Next, the primary outcome measure was based on a single verbal memory and learning test with a ceiling and a floor effect. Using a measure with a more comprehensive score range may have tracked subtle differences between ages. Thus, our results may not be generalizable to different cognitive functions. Further, our study design is dependent on self-report of smoking and cardiovascular disease and we did not attempt to verify these with medical records. However, previous studies have reported discrepancies between self-reported smoking habits and serum cotinine concentration, a biomarker of nicotine absorption, especially in girls and women, suggesting that more women under-report smoking than do men (66–68). A higher rate of inaccurate smoking status self-report by women in our dataset would have attenuated our results. This suggests that our results may actually underestimate the difference between men and women who smoke. Additionally, we were unable to assess a dose-response of smoking since we only asked for smoking status and did not collect requisite smoking history information in order to calculate pack years. Finally, the cross-sectional design of this study does not allow a causal conclusion and a longitudinal study design should follow to verify our results and assess a causal interpretation.

Despite these limitations, there are several advantages of using a large web–based study cohort. Our cohort includes a wide range of adults aged 18 - 85, which allowed us to assess the relationship between smoking and cardiovascular disease and verbal memory in the broadest single study age range used to date. In addition, the MindCrowd cohort has a considerable number of variables assessed, which enabled us to control for many potential confounding variables in addition to conducting propensity matching due to the cohort size. Both regression and propensity analyses indicate that the independent effect of smoking over and above other health factors on verbal memory performance is robust in women. Another advantage of using a web-based study design is the ability to use an identical study wide protocol as opposed to attempting to harmonize protocols and data across sites, which is often required when combining smaller, local cohorts. Furthermore, this cohort is geographically diverse with participants in both rural and urban settings, which allows a higher degree of generalizability. In addition to smoking status, we also explored the synergistic effects of cardiovascular disease on verbal learning and memory using a composite. Composites have the advantage of weighing multiple risks with a single summary variable and can be more sensitive and robust than single variables (69). Therefore, composite scores may be more biologically relevant and have advantages in both clinical practice and cardiovascular research.

In summary, we report that sex moderates the relationship between smoking status and verbal learning and memory performance based on results from the largest study to date. Furthermore, we report down sampling tests that suggest the minimum sample required to dependably detect this interaction 50% of the time is 10,000. Based on these results, prior study sample sizes may have produced less reliable results due to small sample sizes. Our results highlight the importance of investigating sex as a variable of interest in understanding environmental influences on verbal learning and memory performance across development and aging. The results suggest smoking may impact women’s cognitive health to a greater degree than men.

## Data Availability

Data will be made available in a public repository (e.g. Dryad).

## Authors’ contributions

C.R. Lewis: literature search, writing, and data interpretation

J.S. Talboom: data collection, data interpretation, and writing

M.D. De Both: figures, data collection, and data analysis

A.M. Schmidt: literature search, and figures

M.A. Naymik: figures, data collection, and data analysis

A.K. Håberg: writing

T. Rundek: writing

B.E. Levin: writing

S. Hoscheidt: writing

Y. Bolla: writing

R. D. Brinton: writing

M. Hay: writing

C.A. Barnes: writing

E. Glisky: writing

L. Ryan: writing

M.J. Huentelman: study design, data collection, data interpretation, and writing

## Site design, testing, and administrative support

The MindCrowd site was built by The Lavidge Company (Scottsdale, Arizona) and we acknowledge their support through the donation of significant *pro bono* employee hours and resources. We also acknowledge the administrative support of Kara Karaniuk, Bethine Moore, and Danielle Metz (for IRB and CRC activities); Galen Perry, Jeffrey Watkins, Steve Yozwiak, and Murphy Raine (for marketing and communications activities).

## Participant recruitment

The Alzheimer’s Association TrialMatch is recognized for directing participants to MindCrowd. The Alzheimer’s Prevention Initiative at Banner Alzheimer’s Institute (Phoenix, Arizona) also helped with recruitment, and we acknowledge their support as well, including the Program Director Jessica Langbaum. Recruitment via social media was greatly facilitated by the efforts of Target Latino (Havi Goffan, CEO, Atlanta, Georgia). We would also like to thank our MindCrowd Facebook friends, Twitter followers, Redditors, and other social media participants.

